# Seroprevalence of *Toxoplasma gondii* Among Pregnant Women Attending Hospital Universitario Hernando Moncaleano Perdomo in Neiva, Huila, Colombia

**DOI:** 10.64898/2025.12.09.25341876

**Authors:** Frank Barreiro Sanchez, Leidy Yovana Urbano, Juan Sebastián Cuellar, Roberto Losada, Juan Sebastián Henao, Henry Ostos Alfonso, Jairo Antonio Rodriguez

## Abstract

**Background:** Toxoplasmosis is a zoonotic disease caused by Toxoplasma gondii that can have serious consequences in pregnant women and their fetuses, including spontaneous abortion, fetal death, and congenital malformations. In Colombia, congenital toxoplasmosis represents a significant public health concern, with 2-10 cases per 1,000 live births annually. More than half of Colombian pregnant women (50-60%) have anti-Toxoplasma antibodies, indicating high parasite exposure and circulation. Despite documented disease burden in Colombia, epidemiological data from specific regions remain scarce, particularly in the department of Huila.

**Methodology and Principal Findings:** We conducted a descriptive retrospective study of 984 pregnant women attending Hospital Universitario Hernando Moncaleano Perdomo in Neiva, Huila, Colombia, from June to December 2021. Demographic data and serological testing results for IgG and IgM antibodies against T. gondii were collected from medical records. The seroprevalence of IgG was 54% (532/984), while IgM seroprevalence was 3% (30/984). A significant association was found between parity and IgG seropositivity (p = 0.043), with multiparous women showing higher seroprevalence (56.7%) compared to primiparous women (50.3%). No significant associations were found between maternal age or type of delivery and seroprevalence.

**Conclusions and Significance:** This study presents the first epidemiological investigation of toxoplasmosis in Huila department and highlights the substantial burden of T. gondii infection in this region. Parity emerges as a significant factor associated with infection. These findings underscore the urgent need for region-specific public health interventions, including prenatal screening programs and maternal education strategies, to prevent congenital toxoplasmosis and reduce disease burden in this high-prevalence Colombian region.

**AUTHOR SUMMARY:** Toxoplasmosis is a parasitic infection affecting millions worldwide, particularly in warm tropical regions like Colombia. While most healthy people have no symptoms, the infection poses serious threats to pregnant women and unborn babies. If a pregnant woman becomes infected for the first time during pregnancy, the parasite can cross the placenta and cause severe birth defects, miscarriage, or fetal death.

In Colombia, more than half of all pregnant women have been exposed to this parasite, yet little was known about its prevalence in specific regions. We examined 984 pregnant women at a hospital in Neiva, Huila, Colombia, to determine exposure rates. We found that 54% had been infected at some point. Notably, women who had been pregnant before were more likely to carry antibodies than first-time mothers, suggesting exposure increases with pregnancy experience.

These findings highlight the urgent need for better screening and preventive strategies for pregnant women in this region to protect unborn babies from this serious infection.

## INTRODUCTION

Toxoplasmosis is a disease that originates from the presence of the parasite Toxoplasma gondii (*T. gondii*) [1]. Toxoplasmosis was ranked fourth by the Food and Agriculture Organization of the United Nations (FAO) and the World Health Organization (WHO) among the 24 foodborne pathogens that pose the greatest health risks [2]. In approximately 80% of cases, infection is asymptomatic in individuals with a healthy immune system [1]. However, in persons with compromised immune systems, pregnant women without antibodies, and newborns, clinical symptoms can be severe [3]. Human infection occurs when oocysts (sporozoites) present in water, vegetables, or fruits contaminated with cat feces are ingested, or when cysts (bradyzoites) present in raw or undercooked meat are consumed [4]. Additionally, other transmission methods include blood transfusion, laboratory accidents, or organ transplants. There is also the possibility of vertical transmission, that is, from mother to child, which probably occurs through tachyzoites in maternal blood that cross the placenta, resulting in congenital toxoplasmosis [5]. Toxoplasmosis is ubiquitous and spreads throughout the world, infecting both humans and warm-blooded animals [6]. It is estimated that approximately one-third of the global population is infected with T. gondii. The highest prevalence rates are found in humid tropics, while the lowest are observed in cold regions [7,8]. Regional variations in seroprevalence are influenced by climatic conditions that may or may not favor the survival of oocysts, as well as by cultural and socioeconomic factors of each subpopulation [8].

The effects of toxoplasmosis on the health of the mother and newborn require special attention [9]. In Colombia, epidemiological surveillance of the disease focuses on preventing significant clinical manifestations: congenital toxoplasmosis and reactivation in immunocompromised patients. However, both clinical forms continue to represent an important health problem for the country. On average, 2 to 10 cases of congenital infection are registered per 1,000 live births per year. This rate, comparatively high in relation to other countries where the rate is 2 to 10 cases per 10,000 live births, underscores the importance of effectively addressing cerebral toxoplasmosis in HIV patients. This persistent challenge represents a significant enigma for the health system [10,11].

In Colombia, more than half of pregnant women (50-60%) have anti-Toxoplasma antibodies, which indicates high exposure and circulation of the parasite in the country, and between 0.6 to 3% of pregnant women acquire the infection during pregnancy. This risk is higher in adolescents (seroconversion 1.5%) than in pregnant women older than 35 years (seroconversion 0.7%) [12]. Therefore, it is crucial to understand the disease burden in the region. Given that there are no previous studies in this area, it is even more important to investigate and understand the local situation. Previous studies have emphasized the importance of assessing the prevalence of infection in different regions of Colombia, with the purpose of facilitating early detection, precise diagnosis, and timely treatment to prevent serious complications in both pregnant women and fetuses. Likewise, determination of parasite circulation in the region can be fundamental to strengthening disease prevention and control programs [13].

## MATERIALS AND METHODS

### Study Area

This descriptive retrospective study was conducted in Neiva, located in southwestern Colombia, in the department of Huila. Neiva is situated in a strategic geographic location of 2°55′39″ N, 75°17′15″ West[14]. It is a city of regional importance with an estimated population of 357,392 inhabitants (according to 2023 data). The climate in Neiva is predominantly tropical, with warm temperatures throughout the year and a rainy season. The city is located in a specific bioclimatic zone.

### Study Population

This retrospective study included 984 pregnant women who attended Hospital Universitario Hernando Moncaleano Perdomo in the city of Neiva, Huila, Colombia, whose pregnancies came to term during the period from June to December 2021, meeting the established inclusion and exclusion criteria. Demographic data and medical history, such as the presence of IgG and IgM antibodies, maternal age, parity, and type of delivery, were collected from the hospital’s medical records. It is important to note that personal identifiers were not included in the data collection forms, thus safeguarding the privacy and confidentiality of the participants.

### Inclusion/Exclusion Criteria

Of 984 pregnant women treated for pregnancy completion, those with serological diagnosis of *T. gondii* infection during pregnancy all met the inclusion criteria. All pregnant women who underwent serological testing for toxoplasmosis during pregnancy were included. Exclusion criteria included all pregnant women with incomplete information (pregnant women without information on age, parity, and abortion history).

### Statistical Analysis

Data collected in the collection instrument were tabulated in a database designed in Excel, then exported to IBM SPSS Statistics Version 25 statistical software.

For qualitative variables, relative and absolute frequencies were established; chi-square association tests were performed to determine the existence or absence of relationship between qualitative variables; tables and graphs of their frequencies and percentages were made. Statistical significance was established at a p-value < 0.05.

## RESULTS

### Demographic Characteristics of Pregnant Women

In this retrospective study, 984 pregnant women aged 12 to 44 years were included in the analysis and had complete information on IgG and IgM antibodies, maternal age, parity, and type of delivery. The mean age was 25.8 ± 6.5 years. Most pregnant women were in the 18 to 25 years age group. We observed that the majority of women examined were multiparous 64.7% (637/984), primiparous 35.3% (347/984) (Table 1).

**TABLE 1.** SOCIODEMOGRAPHIC CHARACTERISTICS OF PREGNANT WOMEN.

| Characteristic | Numbers | (%) | 95% CI* |
| --- | --- | --- | --- |
| **Mean age (years):** $25.8 \pm 6.5$ | | | |
| <18 | 87 | 8.8% | 15.49 - 15.99 |
| 18 - 35 | 799 | 81.2% | 25.06 - 27.70 |
| >35 | 98 | 10.0% | 37.94 - 38.77 |
| **Total** | **984** | **100%** |  |
| **Parity** |  |  |  |
| Primiparous (1 delivery) | 347 | 35.3% |  |
| Multiparous (>2 deliveries) | 637 | 64.7% |  |
| **Type of Delivery** |  |  |  |
| Cesarean | 472 | 48% | 26.14 - 27.27 |
| Vaginal | 512 | 52% | 24.42 - 25.59 |
| **Total** | **984** |  |  |

\* CI = Confidence interval

### Serological Status of Pregnant Women

According to serological status for *T. gondii* infection, among 984 pregnant women, results showed that the seroprevalence of IgG specific for T. gondii was 54% (532/984) and 46% (452/984) were seronegative for IgG antibodies; furthermore, the percentage of IgM antibodies positive for *T. gondii* was 3% (30/984) and negative 97% (954/984).

### Risk Factors Associated with *T. gondii* Infection

Table 2 provides a detailed analysis of toxoplasmosis seroprevalence in pregnant women, considering age, parity, and type of delivery. It was observed that there are no significant variations in seroprevalence according to age, with similar percentages in the groups <18 years (54%), 18-35 years (53.9%), and >35 years (58.2%). Chi-square analysis (χ^2^ = 2.075, p = 0.722) confirms the lack of statistically significant association between age and toxoplasmosis seroprevalence. However, a significant difference is evident in seroprevalence between primiparous (50.3%) and multiparous (56.7%) women (χ^2^ = 6.294, p = 0.043). Regarding type of delivery, no significant differences in seroprevalence are found between cesarean deliveries (55.1%) and vaginal deliveries (53.7%) (χ^2^ = 1.501, p = 0.472). In summary, parity shows a significant association with toxoplasmosis seroprevalence, while age and type of delivery do not present significant associations.

**TABLE 2.** TOXOPLASMOSIS SEROPREVALENCE ACCORDING TO AGE, PARITY, AND TYPE OF DELIVERY AMONG PREGNANT WOMEN.

| Variable | Seroprevalence of Toxoplasma | | $\chi^2$ | p-value |
| --- | --- | --- | --- | --- |
|  | Seropositivity, n (%) | Seronegativity, n (%) |  |  |
| Age |  |  |  |  |
| <18 | 47 (54%) | 40 (46%) | 2.075 | 0.722 |
| 18 - 35 | 431 (53.9%) | 368 (46.1%) |  |  |
| >35 | 57 (58.2%) | 41 (41.8%) |  |  |
| Parity |  |  |  |  |
| **Primiparous (1 delivery)** | **180 (50.3%)** | **178 (49.7%)** | **6.294** | **0.043\*** |
| **Multiparous (>2 deliveries)** | **355 (56.7%)** | **271 (43.3%)** |  |  |
| **Type of Delivery** |  |  |  |  |
| **Cesarean** | **260 (55.1%)** | **212 (44.9%)** | **1.501** | **0.472** |
| **Vaginal** | **275 (53.7%)** | **237 (46.3%)** |  |  |

\* Statistically significant

## DISCUSSION

As evidenced in multiple studies, the prevalence of *Toxoplasma gondii* varies according to geographic location, the strains involved, and associated risk factors; therefore, great disparity can be found in its percentage of presentation ranging from 18% to 90%. Bartolomé Álvarez presents an analysis of studies conducted in groups of women of childbearing age and pregnant women between 2000 and 2008, showing 41% in Poland, 26% in Sweden, 22% in Italy, 20% in Greece, 14% in Stockholm, and 9% in the United Kingdom [15]. In a study by Wam and collaborators, a high prevalence of the disease (88%) is evident in a population of women of childbearing age in northwestern Cameroon [16]. In our territory, the study by Castro and collaborators showed a seroprevalence of antibodies of 52% in a population of pregnant women in Villavicencio [17], whose results correlate with those evidenced in this study, which showed a prevalence of 54%.

Regarding the detection of IgM antibodies for toxoplasmosis in this study, positivity was observed in 30 cases, representing approximately 3% of the studied population. This figure contrasts with the trend observed globally, where the prevalence of positive IgM is around 1.5%, as demonstrated in the STRAT study conducted in France [18], and as mentioned in the guidelines of the Spanish Society of Pediatric Infectology [19]. In Brazil, a study conducted in 2011 reported an incidence of 2.3% [20]. At the local level, although studies are limited, one conducted in 2014 in two hospitals in Bogotá recorded an incidence of 6% [21]. This disparity in positivity rates could be attributed to various factors, such as differences in public health practices, environmental exposure to the parasite, and the prevalence of the disease in the studied population.

In this study, the seroprevalence of IgG antibodies against *T. gondii* in pregnant women was 54%, while in the study conducted in Rabat, Morocco, they reported a seroprevalence of 43% for IgG. Additionally, it was found that 3% of pregnant women in this study were positive for IgM antibodies against *T. gondii*, in comparison with 3.9% reported in Rabat. Regarding risk factors associated with *T. gondii* infection, the parasitic infection was more prevalent in the 18 to 35 years age group, with 80.5%, followed by the age group over 35 years with 10.7%. In contrast, the study in Rabat showed that *T. gondii* infection was more prevalent in women aged 31 to 35 years, with 50.7%. Additionally, multiparous women were observed to be more immune to infection, with 66.5%, while in Rabat, multiparous women had a seroprevalence of 51.2%. Regarding type of delivery, no significant differences were found, with 48.7% cesarean sections and 51.3% vaginal deliveries in this study, while in the Rabat study, no specific data were mentioned regarding type of delivery in relation to *T. gondii* infection [9].

The comparison of the results of this study and the study by Sadegi Hariri et al. on the seroprevalence of *Toxoplasma gondii* in pregnant women in Ardabil, Iran, reveals some notable similarities and differences. Regarding the seroprevalence of IgG antibodies specific for *T. gondii*, it was observed that 54% of pregnant women in the study sample were seropositive, in contrast to 22.1% reported in Ardabil, suggesting a significantly higher prevalence of IgG antibodies in the studied population. Regarding IgM antibodies, 3% seropositivity was identified in the study, while in the Sadegi Hariri et al. study, no IgM antibodies were detected in any of the participants. This disparity could be attributed to several reasons, such as differences in the studied populations, detection methodologies, or environmental and health conditions. When analyzing risk factors associated with *T. gondii* infection, the parasitic infection was more prevalent in women aged 18 to 35 years in the study, while in Ardabil, no significant differences were observed based on age. Regarding parity, the study showed that multiparous women had greater immunity to infection, which was statistically significant, while in Ardabil, no significant association was found between seroprevalence and parity. Regarding type of delivery, both studies found no significant differences in *T. gondii* seropositivity between cesarean and vaginal deliveries, suggesting that the delivery method did not influence exposure to infection in both populations studied. These discrepancies highlight the importance of considering regional and population variations when interpreting the results of epidemiological studies on toxoplasmosis in pregnant women [22].

When comparing the results of seroprevalence of IgG and IgM antibodies specific for *T. gondii* in pregnant women between this study and the study conducted in the province of El Oro, Ecuador, notable differences are observed. While in this study we found a seroprevalence of 54% for IgG and 3% for IgM, the study in El Oro reported a seroprevalence of 16% for IgG and 0.8% for IgM. Additionally, these findings suggest a higher prevalence of infection in women aged 18 to 35 years and in multiparous women, without significant differences in type of delivery. In contrast, the study in El Oro identified a higher prevalence in women aged 26 to 30 years and in women with multiple pregnancies. These discrepancies could be attributed to differences in the characteristics of the studied populations, the methodologies employed, and the specific environmental factors of each region. It is essential to consider these variations when interpreting these results and when discussing the implications of this study in the context of existing literature on Toxoplasma gondii in pregnant women [23].

The results of this study reveal a significant difference in the seroprevalence of IgG antibodies specific for *T. gondii* in pregnant women compared to the study conducted in Quito, Ecuador, which reported a seroprevalence of 16.32%. This disparity suggests important variations in the prevalence of *T. gondii* infection between the studied populations. These variations could be influenced by diverse factors, such as the geographic location of the regions where the studies were conducted, the environmental conditions characteristic of each area, and the particular life habits of pregnant women in each community. Such factors can affect the exposure of pregnant women to the parasite and, consequently, the prevalence of infection. However, both studies identified age and parity as relevant risk factors for *T. gondii* infection. In both investigations, greater seropositivity was observed in the 18 to 35 years age group, and multiparous women showed greater immunity to infection compared to primiparous women. Furthermore, in both this study and the one conducted in Quito, no significant differences were found in seropositivity between cesarean deliveries and vaginal deliveries, suggesting that the type of delivery may not be a determining factor in exposure to *T. gondii* infection during pregnancy in these populations. These similarities and differences highlight the importance of considering the specific characteristics of each population when analyzing the epidemiology and factors associated with *T. gondii* infection in pregnant women [24].

## CONCLUSIONS

In conclusion, the results of this study and its comparison with previous research underscore the diversity in the prevalence of *Toxoplasma gondii* among different regions and groups of pregnant women. The identification of risk factors such as age, parity, and type of delivery highlights the importance of addressing the prevention and management of this infection in a personalized manner according to the demographic characteristics of each population. The need for additional research is suggested to better understand the mechanisms of transmission and the clinical implications of *T. gondii* infection during pregnancy. These findings emphasize the need to implement public health strategies adapted to the particularities of each regional context.

## Data Availability

All data supporting the findings of this study are available from the corresponding author (Frank Barreiro Sánchez) upon reasonable request. Patient-level data cannot be made publicly available due to ethical considerations and institutional data protection policies that safeguard participant confidentiality. However, interested researchers may contact the corresponding author to request access to anonymized data, subject to approval from the Institutional Review Board at Hospital Universitario Hernando Moncaleano Perdomo and the authors.

